# Weakly-Supervised Gated Attention Multiple Instance Learning for Multi-Center Cervical Cytology Slide Triage

**DOI:** 10.64898/2026.09.29.26364279

**Authors:** Simeon Mayala, Sang’udi Sang’udi, Deogratias Mzurikwao, Emmanuel Suluba

## Abstract

Automated triage of whole-slide cervical cytology images remains constrained by the heavy dependence on single-cell bounding boxes or pixel-level annotations, which introduce significant bottlenecks, high expert overhead, and observer variability. To address these challenges, we present a weakly-supervised, detection-free Multiple Instance Learning (MIL) framework powered by a Gated Attention pooling engine that operates exclusively on slide-level predictions. By treating whole-slide images as bags of local instance patches, the dual-branch gated network dynamically assigns non-linear attention weights to highlight isolated dysplastic cells while suppressing benign background and debris, successfully resolving the “needle-in-a-haystack” problem inherent to high-grade lesions. Evaluated across multi-center cohorts including internal datasets (SIPaKMeD, Herlev, and CRIC) and the unannotated, out-of-distribution Mendeley LBC validation cohort processed via an unsupervised marker-controlled watershed pipeline our approach demonstrates robust generalization without requiring dense instance labels. Furthermore, our architectural comparison reveals a key trade-off: lightweight backbones like MobileNetV2 optimize in-distribution multi-center accuracy (90.96% Accuracy, 0.9800 ROC-AUC), whereas higher-capacity models like Xception ensure superior out-of-distribution robustness (80.43% Balanced Accuracy) under domain shifts. Ultimately, this detection-less pipeline provides a scalable and clinically viable solution to automate slide triage and alleviate cytopathologist shortages in resource-constrained environments.

**Author summary:** Automated screening of cervical cytology slides is traditionally hindered by the need for labor-intensive, pixel-level cell annotations. To overcome this limitation, this study introduces a weakly-supervised, detection-free Multiple Instance Learning framework that performs slide triage using only slide-level diagnostic labels. By employing a Gated Attention mechanism, the model dynamically focuses on informative cellular regions while ignoring background debris, successfully addressing the challenge of identifying rare dysplastic cells across gigapixel images. Evaluations across multi-center datasets demonstrate that lightweight models like MobileNetV2 achieve high accuracy on in-distribution data, whereas deeper architectures like Xception provide superior robustness against out-of-distribution variations. Ultimately, this approach offers a scalable, annotation-free triage tool designed to assist cytopathologists and improve screening workflows in resource-constrained environments.

## 1 Introduction

Although cervical cancer is largely preventable through timely screening, it remains a major driver of female cancer mortality worldwide [1]. Frontline screening modalities such as Pap smears and liquid-based cytology (LBC) produce high-resolution cellular images that are categorized under the Bethesda System into classes including Negative for Intraepithelial Lesion or Malignancy (NILM), Low-Grade Squamous Intraepithelial Lesion (LSIL), and High-Grade Squamous Intraepithelial Lesion (HSIL). However, manual visual inspection of cytology images is labor-intensive, subject to inter-observer variability, and prone to high false-negative rates. The sheer volume of cells per slide creates a severe diagnostic bottleneck, particularly in low- and middle-income countries (LMICs) with a critical shortage of trained cytopathologists.

To alleviate this diagnostic burden, computer-aided automated screening systems powered by deep learning have emerged as a promising solution. However, developing automated deep learning systems for whole-slide cytology image analysis faces three primary operational and algorithmic bottlenecks. First, conventional fully-supervised methods suffer from a granular annotation bottleneck, as pixel- or box-level single-cell annotations across whole slides demand hundreds of expert pathologist hours, making large-scale dataset curation impractical [2, 3]. Second, cytology slides exhibit extreme bag imbalance and diagnostic sparsity; pathological dysplastic cells (e.g., HSIL) often constitute under 1% of the thousands of cells present, causing standard weakly-supervised aggregation methods such as mean-pooling to dilute rare diagnostic signals, while max-pooling leads to severe gradient sparsity [4, 5]. Finally, multi-center deployment is hampered by severe inter-site domain shifts resulting from variations in staining protocols (e.g., Papanicolaou vs. H&E), slide scanner hardware, optical resolution, and preparation techniques across clinical cohorts [6, 7]

To overcome these constraints without requiring dense cell-level labels, recent workflows partition whole-slide images (WSIs) into patches, casting slide-level triage into a Multiple Instance Learning (MIL) framework. Following recent detection-free paradigms in cervical cytopathology whole-slide image (WSI) classification [8], we frame slide-level triage as a weakly-supervised problem. To aggregate patch-level embeddings produced by MobileNetV2 [9], Xception [10] and ConvNeXt-Tiny [11] without losing local morphological nuance, we adopt a Gated Attention Multiple Instance Learning mechanism [4, 12]. This allows the network to automatically assign higher attention weights to key diagnostic cellular clusters (e.g., HSIL, SCC, or ASC-H) while suppressing background debris.

## 2 Materials and methods

### 2.1 Material: Multi-Center Cervical Cytology Corpus

To establish a comprehensive and domain-diverse benchmark for automated cervical cell analysis, we integrated four distinct cytology datasets. This multi-center corpus spans both conventional Pap smears and liquid-based cytology (LBC) preparations:

- **SIPaKMeD Dataset:** A benchmark resource for single-cell classification comprising 4,049 isolated cell images manually cropped from 966 cluster images captured via an optical microscope [13]. It is categorized into five distinct morphological classes (Superficial-Intermediate, Parabasal, Koilocytotic, Dyskeratotic, and Metaplastic) with expert-outlined nuclear and cytoplasmic boundaries.
- **CRIC Cytology Dataset:** Consists of 400 high-resolution whole-slide conventional Pap smear images containing 11,534 expert-annotated cells categorized into six Bethesda system classes [14]. Using the cell-level bounding coordinates provided in the annotations, a dynamic cropping mechanism extracts patch regions centered around each individual annotated cell.
- **Herlev Dataset:** Comprises 917 single-cell images extracted from Pap smear slides across seven cyto-pathological categories: three normal cell types (superficial squamous, intermediate squamous, and columnar epithelium) and four abnormal dysplastic stages (mild, moderate, severe dysplasia, and carcinoma in situ) [15]. Each image includes expert diagnostic labels and manually segmented cytoplasm and nuclear ground-truth masks.
- **Mendeley Liquid-Based Cytology (LBC) Dataset:** Serving as an external out-of-distribution validation benchmark to evaluate model generalization on real-world hospital data, this corpus consists of 963 Pap smear images collected from 460 patients across public hospital O&G departments, captured at 40×magnification using a Leica ICC50 HD microscope [16].

### 2.2 Methodology

This section details the weak-supervision Multiple Instance Learning (MIL) framework designed for slide-level triage across multicenter cervical cytology cohorts. The pipeline spans four interdependent phases: (1) Data Preparation and Bag Construction with Stochastic Instance Subsampling, (2) Exploratory Feature Extractor Paradigms, (3) Dynamic Feature Aggregation via Gated Attention MIL Architecture, and (4) Optimization Dynamics, Cross-Validation, and Evaluation Metrics.

#### 2.2.1 Data Preparation and Bag Construction with Stochastic Instance Subsampling

Let *S* = {SIPaKMeD, Herlev, CRIC} represent the set of heterogeneous collection sites. We formalize the multicenter cytology corpus as 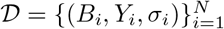 comprising *N* whole-slide image representations, where each slide bag *B*_*i*_ originates from site *σ*_*i*_ ∈*S*.

To maintain realistic biological and patient-level distributions across all modalities, isolated patch and single-cell instances from **SIPaKMeD** and **Herlev** were traced back to their original recorded slide names and grouped into their respective slide bags *B*_*i*_. For the **CRIC** dataset, dynamic cropping was executed around each annotated cell coordinate to isolate cell patches. Prior to bag construction, all extracted patches across all datasets were standardized to a uniform spatial dimension using zero-padding to strictly preserve the underlying cellular aspect ratios and prevent geometric distortion.

Each WSI is thus modeled as an unordered bag *B*_*i*_ containing *K*_*i*_ region-of-interest (ROI) cell patch embeddings:

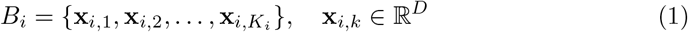

where *D* denotes the feature dimensionality determined by the frozen vision backbone architecture, and *K*_*i*_ reflects the variable cell density of slide *i*. Each slide bag is assigned a global diagnostic categorical label *Y*_*i*_ ∈{0, 1, 2}, mapped directly from international Bethesda System criteria [17]:

- *Y*_*i*_ = 0: **NILM** (Negative for Intraepithelial Lesion or No Malignancy)
- *Y*_*i*_ = 1: **LSIL** (Low-Grade Squamous Intraepithelial Lesion / ASC-US)
- *Y*_*i*_ = 2: **HSIL** (High-Grade Squamous Intraepithelial Lesion / ASC-H / SCC)

Under the weak-supervision MIL paradigm [18], granular patch-level labels *y*_*i,k*_ ∈ {0, 1, 2} remain unobserved during training. While classic MIL posits a deterministic max-pooling relationship *Y*_*i*_ = max_*k*_ *y*_*i,k*_, our framework replaces this non-differentiable operator with a learnable, parameter-efficient attention mechanism over the latent instance subspace [4].

To mitigate out-of-memory (OOM) faults arising from extreme instance variances (*K*_*i*_ ≫ 10^3^) and to regularize against bag-level overfitting, a dynamic stochastic subsampling operator T_sub_ is applied during training passes [19]:

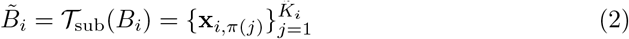

where *π*(·) is a uniform random permutation over indices {1, …, *K*_*i*_}, and the active instance cardinality 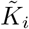 is bounded by:

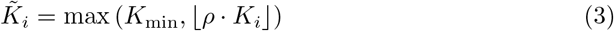

where *ρ* = 0.85 specifies the stochastic instance selection ratio and *K*_min_ = 8 enforces a structural minimum instance threshold. During validation and testing evaluation steps, *T*_sub_ is deactivated (*ρ* = 1.0) to guarantee deterministic bag execution 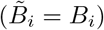.

To handle class imbalance across multicenter diagnostic splits, inverse class frequencies are evaluated over the training split *D*_train_ [20]. Defining 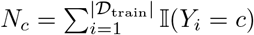 as the total number of slide bags belonging to category *c* ∈ {0, 1, 2}, the class weight vector **w** = [*w*_0_, *w*_1_, *w*_2_]^*T*^ ∈ ℝ^3^ is derived as:

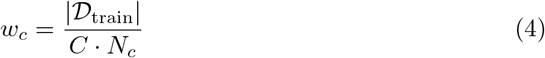

where *C* = 3 represents the total number of diagnostic target categories.

#### 2.2.2 Exploratory Framework: Feature Extractor Architecture Paradigms

To systematically investigate how patch-level representation capacity influences whole-slide cervical cytology classification, all feature extraction backbones (MobileNetV2, Xception, and ConvNeXt-Tiny) were kept strictly frozen throughout training. Pre-trained ImageNet weights were used exclusively as fixed, off-the-shelf feature extractors without backpropagating gradients into the vision backbones.

Each pre-trained backbone maps an input patch image *x*_*i,k*_ to a static embedding vector *h*_*i,k*_ ∈ ℝ^*D*^:

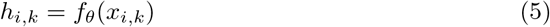

where *D* ∈{1280, 2048, 768}. By freezing *θ* and fixing all downstream aggregation parameters, loss formulations, and hyperparameter schedules, any observed performance variations directly isolate the intrinsic representation quality and domain transfer capabilities of each backbone’s static feature space.

**Fig 1.**
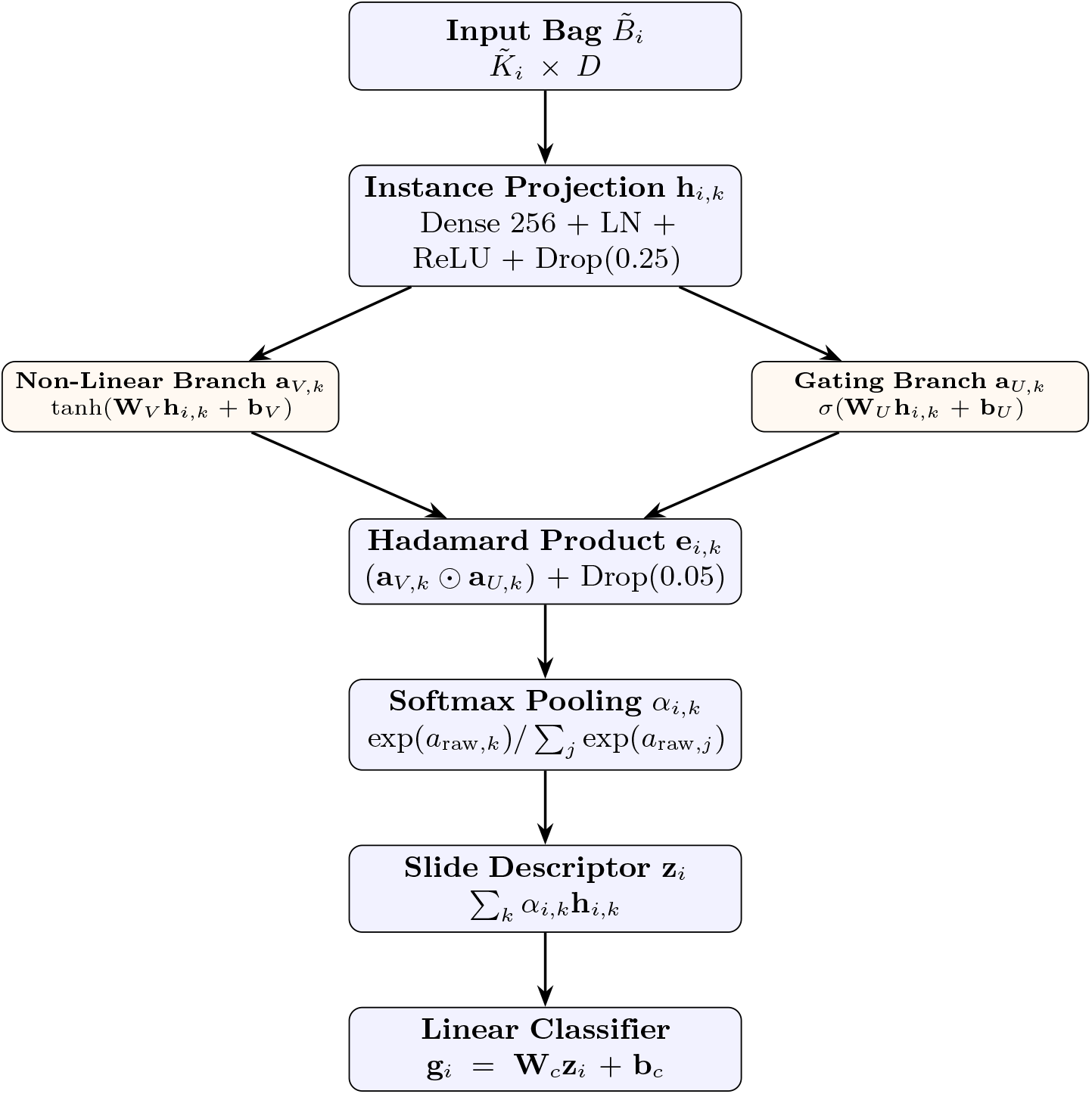
Architectural flow of the Gated Attention MIL Pooling Engine.

#### 2.2.3 Dynamic Feature Aggregation via Gated Attention MIL Architecture

The Gated Attention MIL model transforms an instance collection 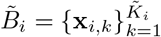 into a fixed-length slide descriptor **z**_*i*_ using non-linear projection, dual-branch gated attention, and permutation-invariant aggregation [2, 4].

Each instance vector **x**_*i,k*_ ∈ ℝ^*D*^ is projected into a baseline latent subspace of dimension *M* = 256 via an encoder module **f**_*θ*_ : ℝ^*D*^ → ℝ^*M*^ :

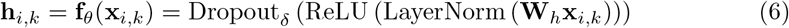

where **W**_*h*_ ∈ ℝ^*M ×D*^, LayerNorm(·) stabilizes feature distributions across sites [21], and Dropout_*δ*_ (·) applies stochastic zeroing at rate *δ* = 0.25 [22].

To capture non-linear morphological variations, a dual-branch gated network targets an intermediate attention dimension *L* = 128 [4, 23]:

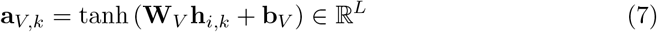

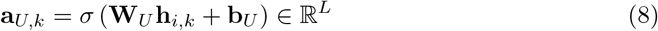

where **W**_*V*_, **W**_*U*_ ∈ ℝ^*L×M*^, **b**_*V*_, **b**_*U*_ ∈ ℝ^*L*^, and *σ*(·) denotes the element-wise logistic sigmoid. Element-wise multiplicative gating yields combined representation **e**_*i,k*_:

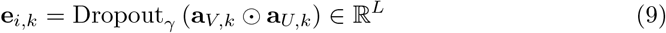

where ⊙ is the Hadamard product and *γ* = 0.05 restricts single-instance dominance. Unnormalized attention weights *a*_raw,*k*_ are generated via linear projection onto vector **w**_*a*_ ∈ ℝ^*L*^:

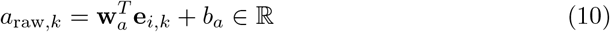

Permutation-invariant pooling normalizes instance importance scores across bag instances [4, 24]:

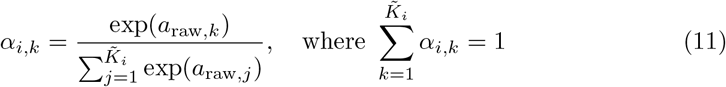

The unified slide representation **z**_*i*_ ∈ ℝ^*M*^ and final logits **g**_*i*_ ∈ ℝ^*C*^ are derived as:

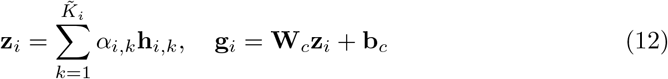

where **W**_*c*_∈ℝ^*C×M*^ and **b**_*c*_∈ℝ^*C*^.

For loss evaluation, ground-truth targets are regularized using Label Smoothing (*ϵ* = 0.03) [25]:

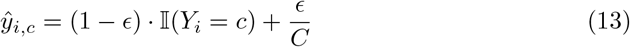

The weighted smoothed cross-entropy loss L_MIL_ for bag *i* is formulated as:

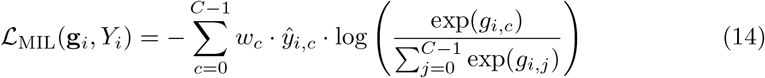

#### 2.2.4 Optimization Dynamics and Evaluation Framework

To maintain stable training on single GPU setups, physical batch size is restricted to *B* = 1 with effective batch size *M*_accum_ = 8 via gradient accumulation. Trainable parameters **Θ** are updated using AdamW (*λ* = 1× 10^*−*4^) [26] following *L*_2_-norm gradient clipping (*τ* = 1.0) [27]. The learning rate follows a Cosine Annealing schedule decaying from *η*_max_ = 3 ×10^*−*4^ to *η*_min_ = 1 ×10^*−*6^ over *E*_max_ = 20 epochs [28]. Checkpoints are selected via early stopping (*P* = 4) on composite metric 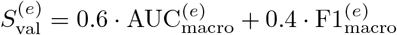.

Robustness is evaluated using a stratified 5-fold cross-validation scheme [29]. Out-of-fold inference vector **P**_OOF_ ∈ℝ^*N ×C*^ aggregates predictions across folds for computing macro-averaged AUC, Precision, Recall, and F1-score via One-vs-Rest (OvR) binarization [30].

### 2.3 Results

#### 2.3.1 Performance Evaluation on Held-Out Test Partitions from Consolidated SIPaKMeD, Herlev, and CRIC Cohorts across Different Backbones

To evaluate model performance on in-distribution multi-center data, we benchmarked the weak-supervision Gated Attention MIL framework across three feature extraction backbones: Xception (*D* = 2048), MobileNetV2 (*D* = 1280), and ConvNeXt-Tiny (*D* = 768). Models were evaluated using both 5-fold cross-validation and an isolated holdout test set partitioned directly from the consolidated dataset comprising the SIPaKMeD, Herlev, and CRIC cohorts. Quantitative performance across all metrics is detailed in Tables 1–4.

**Table 1.** Comparative evaluation of CNN backbones across 5-fold cross-validation and isolated multi-center holdout test set partitioned from SIPaKMeD, Herlev, and CRIC.

| Model | 5-Fold Cross-Validation |  |  | Holdout Test Set |  |  |
| --- | --- | --- | --- | --- | --- | --- |
|  | Macro F1 | Macro AUC | OOF Acc | Accuracy | Macro F1 | ROC-AUC |
| Xception | $0.8552 \pm 0.0119$ | $0.9549 \pm 0.0040$ | 0.8561 | 0.8717 | 0.8699 | 0.9678 |
| MobileNetV2 | $0.8739 \pm 0.0082$ | $0.9642 \pm 0.0068$ | 0.8747 | **0.9096** | **0.9085** | **0.9800** |
| ConvNeXt-Tiny | **$0.8848 \pm 0.0166$** | **$0.9667 \pm 0.0060$** | **0.8860** | 0.8980 | 0.8966 | 0.9764 |

**Table 2.** Isolated multi-center test set per-class F1-score performance on partitioned SIPaKMeD, Herlev, and CRIC data.

| Model | Normal / NILM | LSIL / ASC-US | HSIL / Carcinoma |
| --- | --- | --- | --- |
| Xception | 0.9129 | 0.8374 | 0.8595 |
| MobileNetV2 | **0.9372** | **0.8867** | **0.9016** |
| ConvNeXt-Tiny | 0.9367 | 0.8670 | 0.8862 |

**Table 3.**
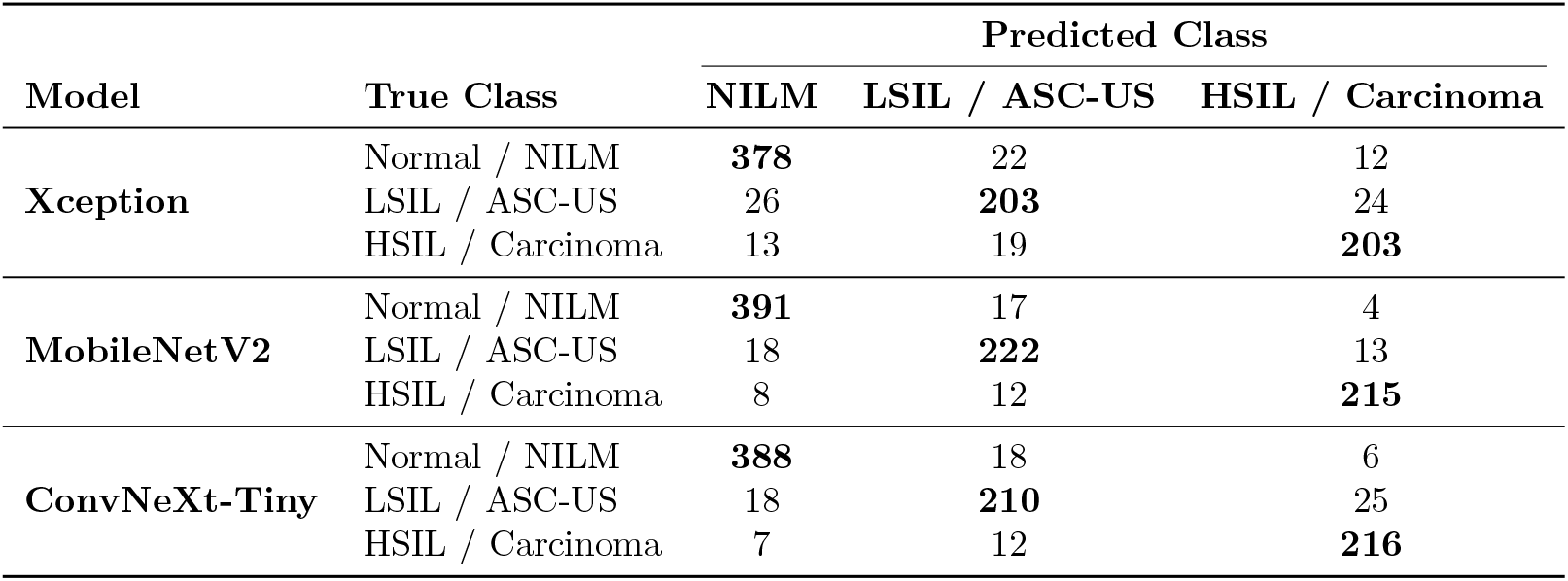
Holdout test set confusion matrices across evaluated architectures on partitioned multi-center data.

| Model | True Class | Predicted Class |  |  |
| --- | --- | --- | --- | --- |
|  |  | NILM | LSIL / ASC-US | HSIL / Carcinoma |
| Xception | Normal / NILM | **378** | 22 | 12 |
|  | LSIL / ASC-US | 26 | **203** | 24 |
|  | HSIL / Carcinoma | 13 | 19 | **203** |
| MobileNetV2 | Normal / NILM | **391** | 17 | 4 |
|  | LSIL / ASC-US | 18 | **222** | 13 |
|  | HSIL / Carcinoma | 8 | 12 | **215** |
| ConvNeXt-Tiny | Normal / NILM | **388** | 18 | 6 |
|  | LSIL / ASC-US | 18 | **210** | 25 |
|  | HSIL / Carcinoma | 7 | 12 | **216** |

**Table 4.** Detailed holdout test set classification metrics on partitioned multi-center data.

| Model | Class / Metric | Precision | Recall | F1-Score | Support |
| --- | --- | --- | --- | --- | --- |
| Xception | Normal / NILM | 0.9064 | 0.9175 | 0.9129 | 412 |
|  | LSIL / ASC-US | 0.8320 | 0.8024 | 0.8374 | 253 |
|  | HSIL / Carcinoma | 0.8494 | 0.8638 | 0.8595 | 235 |
|  | Accuracy | – | – | 0.8717 | 900 |
|  | Macro Avg | 0.8626 | 0.8612 | 0.8699 | 900 |
|  | Weighted Avg | 0.8706 | 0.8717 | 0.8718 | 900 |
| MobileNetV2 | Normal / NILM | 0.9376 | 0.9489 | **0.9372** | 412 |
|  | LSIL / ASC-US | 0.8845 | 0.8775 | **0.8867** | 253 |
|  | HSIL / Carcinoma | 0.8884 | 0.9149 | **0.9016** | 235 |
|  | Accuracy | – | – | **0.9096** | 900 |
|  | Macro Avg | **0.9035** | **0.9138** | **0.9085** | 900 |
|  | Weighted Avg | **0.9098** | **0.9096** | **0.9092** | 900 |
| ConvNeXt-Tiny | Normal / NILM | **0.9395** | 0.9417 | 0.9367 | 412 |
|  | LSIL / ASC-US | 0.8750 | 0.8300 | 0.8670 | 253 |
|  | HSIL / Carcinoma | 0.8745 | **0.9191** | 0.8862 | 235 |
|  | Accuracy | – | – | 0.8980 | 900 |
|  | Macro Avg | 0.8963 | 0.8969 | 0.8966 | 900 |
|  | Weighted Avg | 0.8972 | 0.8980 | 0.8973 | 900 |

##### Comparative Backbone Performance

As shown in Table 1, ConvNeXt-Tiny achieved the highest 5-fold cross-validation metrics on the consolidated training folds, yielding an out-of-fold accuracy of 0.8860 and macro ROC-AUC of 0.9667 ±0.0060. This indicates that modern ConvNet design choices (e.g., 7 ×7 depthwise kernels and LayerNorm) provide strong inductive biases during feature learning.

However, on the isolated holdout test set partitioned from the consolidated SIPaKMeD, Herlev, and CRIC datasets, MobileNetV2 demonstrated superior generalization, outperforming all architectures in classification accuracy (0.9096), Macro F1-score (0.9085), and ROC-AUC (0.9800). MobileNetV2 surpassed ConvNeXt-Tiny by +1.16% in test accuracy and Xception by +3.79%. The moderate feature dimension of MobileNetV2 (*D* = 1280) provided an optimal trade-off between representation capacity and parameter efficiency, mitigating downstream overfitting during gated attention pooling. Conversely, Xception underperformed across benchmarks, suggesting that higher-dimensional embedding spaces (*D* = 2048) can introduce redundant features that hinder precise attention weight estimation (*α*_*i,k*_).

### 2.4 External Out-of-Distribution Validation on Mendeley LBC

To evaluate out-of-distribution (OOD) robustness across all comparative model architectures, we benchmarked our framework on the unseen Mendeley LBC cohort. Crucially, the Mendeley LBC dataset provides only coarse, slide-level diagnostic classifications without any instance-level or cell-wise annotations. To enable weakly-supervised evaluation, an unsupervised candidate-extraction pipeline was implemented to automatically generate patch instances and construct slide-level bags for inference.

Field-of-view images were inverted and thresholded using Otsu’s method to isolate foreground cellular structures. Seed points corresponding to candidate cell centers were identified by locating local maxima on an intensity-weighted distance surface combining normalized pixel intensity (60% weight) and Euclidean distance from object boundaries (40% weight). Marker-controlled watershed segmentation decoupled overlapping cellular structures. Extracted candidate regions were bounded with a 15-pixel spatial padding margin, filtered by area (≥10, 000 px^2^), and standardized with zero-padding to preserve aspect ratios.

To conduct comparative OOD benchmarking, candidate patches extracted from a parent image were mapped to low-dimensional feature representations **h**_*i*_ ∈ℝ^*d*^ using the respective pre-trained, frozen feature extractors specifically, MobileNetV2 (*d* = 1280), Xception (*d* = 2048), and ConvNeXt-Tiny (*d* = 768). For each architecture, extracted instances were aggregated into unannotated slide-level bags ℬ ^OOD^ = {**h**_1_, **h**_2_, …, **h**_*N*_}. This enabled the pre-trained Gated Attention MIL engine corresponding to each backbone to dynamically assign attention weights across candidate cells and predict the overall slide category without requiring cell-wise supervision during evaluation.

#### 2.4.1 5-Fold Model Ensemble Aggregation

To enhance model generalization and mitigate domain shift on unseen OOD validation datasets, predictions across all *M* = 5 models trained on cross-validation folds for a given backbone are ensembled via uniform averaging for each patch *i*:

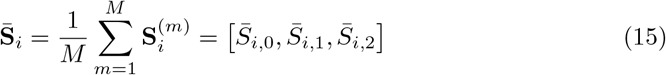

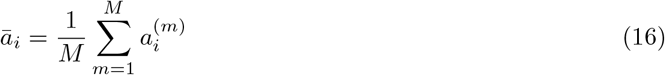

where 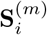 and 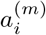 denote the class logits and attention weight predicted by the *m*-th fold model of the corresponding backbone for patch *i*, respectively.

#### 2.4.2 Hierarchical Safety-First Decision Tree

To suppress artifact noise while maintaining absolute sensitivity for high-grade precancerous slides across out-of-distribution domain shifts, an instance-level attention cutoff mask **M** ∈ {0, 1}^*N*^ is applied:

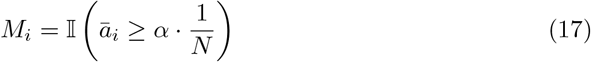

where *α* = 1.0 filters out candidate patches falling below the uniform attention baseline across the slide bag.

Logit margins relative to the normal class (NILM = 0) are calculated for filtered patches for both HSIL 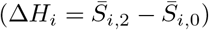 and LSIL 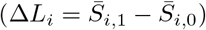. To ensure robustness against variable cellular density in unannotated OOD slides, positive patch ratios are dynamically evaluated against total bag size *N* :

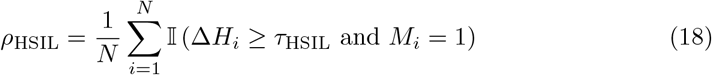

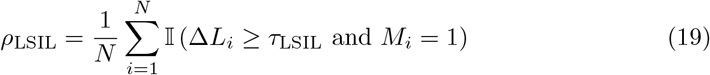

where margin cutoffs are set to *τ*_HSIL_ = 0.0 and *τ*_LSIL_ = 0.5.

Additionally, inter-fold prediction uncertainty is quantified via the normalized Shannon entropy across ensemble probabilities 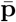:

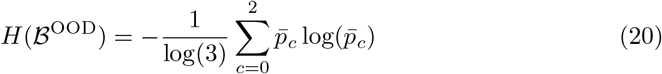

The ultimate slide classification rule *ŷ* ∈0, 1, 2, Manual Review ensures high sensitivity for precancerous lesions while preventing out-of-distribution misclassifications under domain shift:

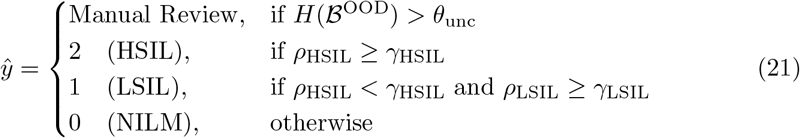

The full out-of-distribution hyperparameter optimization, safety-constrained evaluations, and optimal operating breakdowns across all three feature backbones are comprehensively presented in the following subsections: MobileNetV2 (Tables 5, 6, and 7), Xception (Tables 8, 9, and 10), and ConvNeXt-Tiny (Tables 11, 12, and 13).

**Table 5.** Top 10 Most Balanced Parameter Configurations for MobileNetV2 on Mendeley LBC (Ranked by Macro F1-Score).

| $\tau_{\text{HSIL}}$ | $\tau_{\text{LSIL}}$ | $\alpha_{\text{attn}}$ | $K_{\text{HSIL}}$ | $K_{\text{LSIL}}$ | Macro F1 | Bal. Acc. | Accuracy | NILM Spec. | LSIL Rec. | HSIL Sens. | HSIL Misses | Abn. Sens. |
| --- | --- | --- | --- | --- | --- | --- | --- | --- | --- | --- | --- | --- |
| -3.0 | -3.0 | 1.00 | 10 | 10 | **0.5860** | 0.6139 | **0.6304** | **0.9167** | 0.2727 | 0.7826 | 10 | 0.8088 |
| -1.5 | -1.5 | 1.00 | 5 | 5 | 0.5700 | 0.5661 | **0.6304** | 0.6667 | 0.2273 | 0.9130 | 4 | 0.9265 |
| -3.0 | -1.5 | 1.00 | 10 | 5 | 0.5679 | 0.5608 | 0.5870 | 0.6667 | 0.3636 | 0.9130 | 4 | 0.9265 |
| -1.5 | -3.0 | 1.00 | 10 | 10 | 0.5569 | **0.6422** | 0.5543 | **0.9167** | **0.7273** | 0.7609 | 11 | 0.7941 |
| -1.5 | -3.0 | 1.00 | 5 | 5 | 0.5464 | 0.5257 | 0.5978 | 0.5000 | 0.2727 | **0.9783** | **1** | **0.9853** |
| 3.0 | 0.0 | 0.00 | 1 | 10 | 0.5436 | 0.5454 | 0.5543 | 0.4583 | 0.5909 | 0.8696 | 6 | 0.8971 |
| 3.0 | 0.0 | 0.25 | 1 | 10 | 0.5436 | 0.5454 | 0.5543 | 0.4583 | 0.5909 | 0.8696 | 6 | 0.8971 |
| -3.0 | -3.0 | 1.00 | 10 | 5 | 0.5431 | 0.5204 | 0.5543 | 0.5000 | 0.4091 | **0.9783** | **1** | **0.9853** |
| 3.0 | 0.0 | 0.50 | 1 | 10 | 0.5424 | 0.5527 | 0.5435 | 0.5000 | 0.6364 | 0.8043 | 9 | 0.8529 |
| 3.0 | 3.0 | 0.00 | 1 | 5 | 0.5356 | 0.5277 | 0.5435 | 0.5417 | 0.4545 | 0.6739 | 15 | 0.7206 |

**Table 6.**
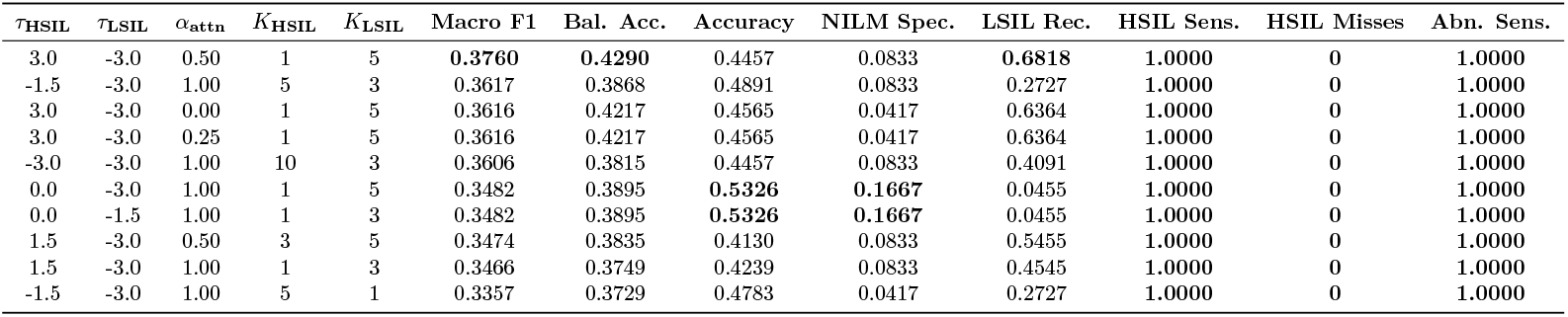
Top 10 Safety-First Parameter Configurations for MobileNetV2 on Mendeley LBC (Constrained to Zero HSIL Misses).

**Table 7.** Detailed Performance Breakdown for MobileNetV2 Under the Optimal Balanced Operating Point (*τ*_HSIL_ = −3.0, *τ*_LSIL_ = −3.0, *α*_attn_ = 1.0, *K*_HSIL_ = 10, *K*_LSIL_ = 10).

|  | Pred NILM | Pred LSIL | Pred HSIL |
| --- | --- | --- | --- |
| True NILM | **22** | 0 | 2 |
| True LSIL | 3 | **6** | 13 |
| True HSIL | 10 | 6 | **30** |

(b) Per-Class Metrics Report
| Class | Precision | Recall | F1-Score | Support |
| --- | --- | --- | --- | --- |
| NILM | 0.6286 | 0.9167 | 0.7458 | 24 |
| LSIL | 0.5000 | 0.2727 | 0.3529 | 22 |
| HSIL | 0.6667 | 0.6522 | 0.6593 | 46 |
| Accuracy | — | — | 0.6304 | 92 |
| Macro Avg | 0.5984 | 0.6139 | **0.5860** | 92 |
| Weighted Avg | 0.6169 | 0.6304 | 0.6086 | 92 |

**Table 8.** Top 10 Most Balanced Parameter Configurations for Xception on Mendeley LBC (Ranked by Macro F1-Score).

| $\tau_{\text{HSIL}}$ | $\tau_{\text{LSIL}}$ | $\alpha_{\text{attn}}$ | $K_{\text{HSIL}}$ | $K_{\text{LSIL}}$ | Macro F1 | Bal. Acc. | Accuracy | NILM Spec. | LSIL Rec. | HSIL Sens. | HSIL Misses | Abn. Sens. |
| --- | --- | --- | --- | --- | --- | --- | --- | --- | --- | --- | --- | --- |
| 0.0 | -3.0 | 1.00 | 3 | 5 | **0.6139** | 0.6113 | 0.6413 | 0.4583 | 0.6364 | 0.9565 | 2 | 0.9706 |
| -1.5 | -3.0 | 1.00 | 5 | 5 | 0.6102 | 0.6040 | **0.6522** | 0.4167 | 0.5909 | 0.9565 | 2 | 0.9706 |
| 0.0 | -3.0 | 1.00 | 5 | 5 | 0.5991 | **0.6376** | 0.5978 | 0.4583 | **0.9545** | 0.9565 | 2 | 0.9706 |
| 0.0 | 0.0 | 1.00 | 3 | 5 | 0.5957 | 0.6037 | 0.6413 | **0.7083** | 0.3636 | 0.8478 | 7 | 0.8088 |
| 0.0 | 0.0 | 0.00 | 10 | 10 | 0.5893 | 0.5930 | 0.6196 | 0.5833 | 0.5000 | 0.8261 | 8 | 0.8235 |
| -3.0 | -3.0 | 1.00 | 10 | 5 | 0.5874 | 0.5902 | 0.5978 | 0.4583 | 0.6818 | 0.9565 | 2 | 0.9706 |
| 0.0 | -1.5 | 0.50 | 5 | 10 | 0.5854 | 0.5724 | 0.6304 | 0.4583 | 0.4545 | 0.9130 | 4 | 0.9265 |
| 0.0 | 0.0 | 0.25 | 10 | 10 | 0.5850 | 0.5937 | 0.6087 | 0.5833 | 0.5455 | 0.7826 | 10 | 0.7941 |
| 3.0 | -3.0 | 1.00 | 1 | 5 | 0.5834 | 0.5921 | 0.6087 | 0.3750 | 0.7273 | **0.9783** | **1** | **0.9853** |
| 0.0 | -1.5 | 0.00 | 10 | 10 | 0.5801 | 0.5842 | 0.6087 | 0.3750 | 0.6818 | **0.9783** | **1** | **0.9853** |

**Table 9.** Top 10 Safety-First Parameter Configurations for Xception on Mendeley LBC (Constrained to Zero HSIL Misses).

| $\tau_{\text{HSIL}}$ | $\tau_{\text{LSIL}}$ | $\alpha_{\text{attn}}$ | $K_{\text{HSIL}}$ | $K_{\text{LSIL}}$ | Macro F1 | Bal. Acc. | Accuracy | NILM Spec. | LSIL Rec. | HSIL Sens. | HSIL Misses | Abn. Sens. |
| --- | --- | --- | --- | --- | --- | --- | --- | --- | --- | --- | --- | --- |
| -1.5 | 0.0 | 1.00 | 5 | 1 | **0.5398** | 0.5484 | **0.6087** | 0.2500 | 0.5909 | **1.0000** | **0** | **1.0000** |
| 0.0 | 0.0 | 1.00 | 3 | 1 | 0.5306 | 0.5418 | 0.5870 | 0.2500 | 0.6364 | **1.0000** | **0** | **1.0000** |
| 3.0 | -1.5 | 1.00 | 1 | 3 | 0.5298 | 0.5504 | 0.5761 | 0.2500 | 0.7273 | **1.0000** | **0** | **1.0000** |
| 1.5 | -1.5 | 0.00 | 3 | 10 | 0.5282 | 0.5149 | 0.5978 | **0.3333** | 0.3636 | **1.0000** | **0** | **1.0000** |
| 1.5 | -1.5 | 0.50 | 3 | 5 | 0.5229 | 0.5346 | 0.5761 | 0.2500 | 0.6364 | **1.0000** | **0** | **1.0000** |
| 1.5 | -1.5 | 0.25 | 3 | 10 | 0.5216 | 0.5077 | 0.5870 | **0.3333** | 0.3636 | **1.0000** | **0** | **1.0000** |
| 0.0 | 0.0 | 1.00 | 5 | 1 | 0.5190 | **0.5682** | 0.5435 | 0.2500 | **0.9545** | **1.0000** | **0** | **1.0000** |
| 3.0 | -3.0 | 1.00 | 1 | 3 | 0.5090 | 0.5365 | 0.5652 | 0.2083 | 0.7273 | **1.0000** | **0** | **1.0000** |
| 3.0 | 0.0 | 1.00 | 1 | 1 | 0.5090 | 0.5365 | 0.5652 | 0.2083 | 0.7273 | **1.0000** | **0** | **1.0000** |
| -3.0 | 0.0 | 1.00 | 10 | 1 | 0.5071 | 0.5208 | 0.5435 | 0.2500 | 0.6818 | **1.0000** | **0** | **1.0000** |

**Table 10.** Detailed Performance Breakdown for Xception Under the Optimal Balanced Operating Point (*τ*_HSIL_ = 0.0, *τ*_LSIL_ = −3.0, *α*_attn_ = 1.0, *K*_HSIL_ = 3, *K*_LSIL_ = 5).

|  | Pred NILM | Pred LSIL | Pred HSIL |
| --- | --- | --- | --- |
| True NILM | 11 | 10 | 3 |
| True LSIL | 0 | 14 | 8 |
| True HSIL | 2 | 10 | 34 |

(b) Per-Class Metrics Report
| Class | Precision | Recall | F1-Score | Support |
| --- | --- | --- | --- | --- |
| NILM | 0.8462 | 0.4583 | 0.5946 | 24 |
| LSIL | 0.4118 | 0.6364 | 0.5000 | 22 |
| HSIL | 0.7556 | 0.7391 | 0.7473 | 46 |
| Accuracy | — | — | 0.6413 | 92 |
| Macro Avg | 0.6712 | 0.6113 | **0.6139** | 92 |
| Weighted Avg | 0.6970 | 0.6413 | 0.6483 | 92 |

**Table 11.**
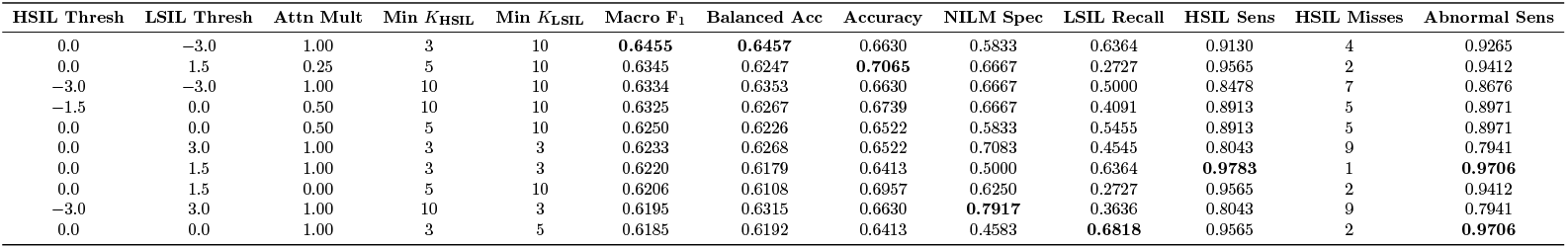
Top 10 Most Balanced Parameter Sets for ConvNeXt-Tiny Gated-Attention MIL (Ranked by Macro F_1_).

**Table 12.** Top 10 Safety-First Parameter Sets for ConvNeXt-Tiny Gated-Attention MIL (Ranked by Minimal HSIL Misses, then Macro F_1_).

| HSIL Thresh | LSIL Thresh | Attn Mult | Min $K_{\text{HSIL}}$ | Min $K_{\text{LSIL}}$ | Macro $F_1$ | Balanced Acc | Accuracy | NILM Spec | LSIL Recall | HSIL Sens | HSIL Misses | Abnormal Sens |
| --- | --- | --- | --- | --- | --- | --- | --- | --- | --- | --- | --- | --- |
| 0.0 | 0.0 | 1.00 | 3 | 3 | **0.5724** | **0.5775** | 0.6087 | 0.3333 | **0.6818** | 1.0000 | **0** | **1.0000** |
| -1.5 | 0.0 | 1.00 | 5 | 3 | 0.5625 | 0.5525 | **0.6304** | 0.3333 | 0.4545 | 1.0000 | **0** | **1.0000** |
| -3.0 | 3.0 | 1.00 | 10 | 1 | 0.5611 | 0.5532 | 0.5978 | 0.3750 | 0.5455 | 1.0000 | **0** | 0.9853 |
| 0.0 | 1.5 | 0.00 | 10 | 3 | 0.5598 | 0.5618 | 0.5652 | **0.4167** | **0.6818** | 1.0000 | **0** | 0.9853 |
| 0.0 | 3.0 | 1.00 | 3 | 1 | 0.5589 | 0.5624 | 0.5978 | 0.3333 | 0.6364 | 1.0000 | **0** | 0.9853 |
| -3.0 | 0.0 | 1.00 | 10 | 3 | 0.5578 | 0.5545 | 0.5978 | 0.3333 | 0.5909 | 1.0000 | **0** | **1.0000** |
| 0.0 | 1.5 | 0.00 | 5 | 3 | 0.5533 | 0.5427 | 0.6413 | 0.3750 | 0.3182 | 1.0000 | **0** | 0.9853 |
| 0.0 | 1.5 | 0.25 | 5 | 3 | 0.5533 | 0.5427 | 0.6413 | 0.3750 | 0.3182 | 1.0000 | **0** | 0.9853 |
| 1.5 | 0.0 | 1.00 | 1 | 5 | 0.5525 | 0.5374 | 0.5978 | 0.3750 | 0.4545 | 1.0000 | **0** | **1.0000** |
| 0.0 | 0.0 | 0.50 | 5 | 3 | 0.5498 | 0.5557 | 0.5978 | 0.2917 | 0.6364 | 1.0000 | **0** | **1.0000** |

**Table 13.** Performance Evaluation and Confusion Matrix for Optimal Balanced ConvNeXt-Tiny MIL Triage Model (HSIL_Thresh_ = 0.0, LSIL_Thresh_ = −3.0, Attn_Mult_ = 1.0, Min *K*_HSIL_ = 3, Min *K*_LSIL_ = 10).

|  | Pred NILM | Pred LSIL | Pred HSIL |
| --- | --- | --- | --- |
| True NILM | 14 | 5 | 5 |
| True LSIL | 1 | 14 | 7 |
| True HSIL | 4 | 9 | 33 |

(b) Per-Class Performance Metrics
| Class | Precision | Recall | F1-Score | Support |
| --- | --- | --- | --- | --- |
| NILM | 0.7368 | 0.5833 | 0.6512 | 24 |
| LSIL | 0.5000 | 0.6364 | 0.5600 | 22 |
| HSIL | 0.7333 | 0.7174 | 0.7253 | 46 |
| Accuracy | — | — | **0.6630** | 92 |
| Macro Avg | 0.6567 | 0.6457 | **0.6455** | 92 |
| Weighted Avg | 0.6785 | 0.6630 | 0.6664 | 92 |

#### 2.4.3 MobileNetV2

The parameter sweep evaluating overall diagnostic performance for MobileNetV2 on the Mendeley LBC cohort is summarized in Table 5. To evaluate clinical triage configurations prioritizing zero false negatives for high-grade lesions, Table 6 details the top safety-first parameter sets under the strict constraint of zero missed HSIL cases.

Under the optimal balanced operating point (*τ*_HSIL_ = −3.0, *τ*_LSIL_ = −3.0, *α*_attn_ = 1.0, *K*_HSIL_ = 10, *K*_LSIL_ = 10), the detailed confusion matrix and per-class classification metrics for MobileNetV2 are reported in Table 7.

#### 2.4.4 Xception

For the Xception feature backbone, Table 8 ranks the top 10 parameter configurations evaluated across overall macro metrics on the Mendeley LBC dataset. The corresponding safety-first hyperparameter combinations achieving complete high-grade sensitivity (0 HSIL misses) are outlined in Table 9. Furthermore, Table 10 presents the subtable pair comprising the 3-class confusion matrix (Table 10a) and the itemized classification metrics (Table 10b) under the primary operating threshold (*τ*_HSIL_ = 0.0, *τ*_LSIL_ = −3.0, *α*_attn_ = 1.0, *K*_HSIL_ = 3, *K*_LSIL_ = 5).

#### 2.4.5 ConvNeXt-Tiny

Finally, the benchmark metrics for ConvNeXt-Tiny on the Mendeley LBC cohort are presented across Tables 11, 12, and 13. Table 11 presents the top 10 balanced parameter choices ranked by macro F_1_-score, while Table 12 delineates the safety-first operating configurations minimizing high-grade false negative errors. Table 13 details the optimal model triage summary, incorporating both the confusion matrix (Table 13a) and the per-class metric breakdown (Table 13b) at the chosen operating thresholds (HSIL_Thresh_ = 0.0, LSIL_Thresh_ = −3.0, Attn_Mult_ = 1.0, Min *K*_HSIL_ = 3, Min *K*_LSIL_ = 10).

## Discussion

The primary contribution of this study lies in demonstrating that high-precision whole-slide cervical cytology triage can be effectively achieved using a weakly-supervised, detection-free Multiple Instance Learning (MIL) framework. Conventional digital cytopathology approaches heavily depend on single-cell bounding boxes or pixel-level annotations to locate dysplastic cells. This micro-level annotation dependency introduces significant bottlenecks, high expert overhead, and susceptibility to observer variability during patch preparation. By shifting the diagnostic paradigm exclusively to slide-level predictions, our framework eliminates the need for manual cell labeling while preserving robust diagnostic accuracy across diverse multicenter cohorts.

### 3.1 Weakly-Supervised Slide-Level Intelligence

Operating strictly under weak bag-level supervision, the Gated Attention engine acts as a dynamic sparse selector. Rather than treating all cropped patches equally, the dual-branch gated network assigns non-linear attention weights to highlight isolated dysplastic cells while suppressing benign cellular background and debris. This design addresses a core challenge in cervical cytology: the “needle-in-a-haystack” problem, where high-grade abnormalities (HSIL) may present in only a fraction of cells across an entire slide. The model learns to identify diagnostic signatures purely from the global Bethesda category of the slide without requiring explicit patch-level supervision.

### 3.2 Trade-Offs in Feature Extraction Backbones

Our comparison across MobileNetV2, ConvNeXt-Tiny, and Xception reveals key architectural trade-offs between parameter efficiency and domain transferability:

- **In-Distribution Optimization:** MobileNetV2 achieved the top overall performance (90.96% Accuracy, 0.9800 ROC-AUC) on internal cohorts. Its lightweight feature dimensionality (*D* = 1280) prevented the Gated Attention mechanism from overfitting to patch embeddings during joint optimization.
- **Out-of-Distribution (OOD) Robustness:** On the unseen Mendeley LBC dataset, Xception yielded superior out-of-distribution balance (80.43% Balanced Accuracy). The deeper, decoupled feature representation of Xception proved better at absorbing the domain shifts inherent to liquid-based preparation variations without relying on fine-grained cell annotations.

### 3.3 Real-World Generalization and Methodological Robustness

A key strength of this framework lies in its structural transition from structured multi-center training cohorts to completely unannotated, real-world clinical deployment. While the training datasets (SIPaKMeD, Herlev, and CRIC) were curated from sources containing initial cell-level annotations to construct representative slide bags, the weakly-supervised MIL paradigm strictly treats granular instance-level labels as unobserved during training. In contrast, the Mendeley LBC validation cohort represents a true out-of-distribution, unannotated hospital environment lacking any pre-existing cell boundaries or ground-truth boxes. For this external evaluation, patches were extracted using an unsupervised marker-controlled watershed segmentation pipeline and assessed solely through coarse slide-level diagnostic labels. This capability highlights the framework’s methodological robustness: while source-domain annotations facilitate structured bag construction, the model successfully generalizes to raw, unannotated hospital data.

## 4 Conclusion

In this study, we formulated whole-slide cervical cytology triage as a weakly-supervised, detection-free Multiple Instance Learning framework powered by a Gated Attention pooling engine. By bridging the gap between benchmark training datasets and unannotated, real-world clinical validation on the Mendeley LBC cohort, our approach successfully bypasses the prohibitive bottleneck of dense single-cell annotations. The evaluation reveals a vital architectural balance: lightweight backbones like MobileNet V2 optimize in-distribution multi-center accuracy, whereas higher-capacity models like Xception ensure robust feature representation under severe out-of-distribution domain shifts. Ultimately, by automating slide triage and prioritizing high-grade precancerous lesions without requiring expert cell-level labeling, this detection-less pipeline provides a scalable, clinically viable solution to alleviate acute cytopathologist shortages in resource-constrained healthcare environments.

## Data Availability

The data set used in this study are freely available online.

https://bids.berkeley.edu/cric-cervix-collection

https://www.kaggle.com/datasets/blank1508/mendeley-lbc-cervical-cancer

https://www.kaggle.com/datasets/prahladmehandiratta/cervical-cancer-largest-dataset-sipakmed

https://www.kaggle.com/datasets/yuvrajsinhachowdhury/herlev-dataset

## Acknowledgments

We are grateful to our colleagues at the ETH Lab for their support and critical review.

## Notes

### Competing Interest Statement

The authors have declared no competing interest.

### Author Declarations

This study did not involve human participants, human tissue, or identifiable human data. Therefore, Institutional Review Board (IRB) approval and informed consent were not required.

